# SARS-CoV-2 Infection of the Placenta

**DOI:** 10.1101/2020.04.30.20083907

**Authors:** Hillary Hosier, Shelli Farhadian, Raffaella A. Morotti, Uma Deshmukh, Alice Lu-Culligan, Katherine H. Campbell, Yuki Yasumoto, Chantal B.F. Vogels, Arnau Casanovas-Massana, Pavithra Vijayakumar, Bertie Geng, Camila D. Odio, John Fournier, Anderson F. Brito, Joseph R. Fauver, Feimei Liu, Tara Alpert, Reshef Tal, Klara Szigeti-Buck, Sudhir Perincheri, Christopher Larsen, Aileen M. Gariepy, Gabriela Aguilar, Kristen L. Fardelmann, Malini Harigopal, Hugh S. Taylor, Christian M. Pettker, Anne L. Wyllie, Charles Dela Cruz, Aaron M. Ring, Nathan D. Grubaugh, Albert I. Ko, Tamas L. Horvath, Akiko Iwasaki, Uma M. Reddy, Heather S. Lipkind

## Abstract

**Background:** The effects of Covid-19 in pregnancy remain relatively unknown. We present a case of second trimester pregnancy with symptomatic Covid-19 complicated by severe preeclampsia and placental abruption.

**Methods:** We analyzed placenta for the presence of SARS-CoV-2 through molecular and immunohistochemical assays and by and electron microscopy, and we measured the maternal antibody response in blood to this infection.

**Results:** SARS-CoV-2 localized predominantly to syncytiotrophoblast cells at the maternal-fetal interface of the placenta. Histological examination of the placenta revealed a dense macrophage infiltrate, but no evidence for vasculopathy typically associated with preeclampsia.

**Conclusion:** This case demonstrates, for the first time, SARS-CoV-2 invasion of the placenta, highlighting the potential for severe morbidity among pregnant women with Covid-19.

## INTRODUCTION

Severe acute respiratory syndrome coronavirus 2 (SARS-CoV-2) is a novel betacoronavirus causing the deadly pandemic of coronavirus disease 2019 (Covid-19). The risks and specific effects of SARS-CoV-2 in pregnant women remain unknown. No adverse outcomes were reported among a small cohort of pregnant women who presented with Covid-19 in the late third trimester of pregnancy in Wuhan, China^1^. However, little is known about maternal and neonatal outcomes as a result of infection in the first and second trimesters of pregnancy.

Hypertensive disorders of pregnancy (HDP) complicate 2-8% of pregnancies and rarely occur in the second trimester^2^. HDP in women with Covid-19 have been noted within limited case reports from China and New York City^3,4^. At the same time, a subset of non-pregnant patients with Covid-19 have demonstrated significant abnormalities of liver enzymes as well as coagulopathy^5,6^. These laboratory abnormalities significantly overlap with findings seen in severe preeclampsia, a subset of HDP, defined by associated findings of hemolysis, elevated liver enzymes, and low platelets as well as proteinuria and elevated blood pressures. This leads to a diagnostic dilemma when faced with pregnant patients with Covid-19, hypertension, and coagulopathy.

We present a case of a woman with Covid-19 in the second trimester of pregnancy with severe hypertension, coagulopathy and preeclampsia. This case highlights the association between Covid-19 and HDP, and demonstrates clear SARS-CoV-2 invasion of the placenta, with associated placental inflammation distinct from typical preeclampsia.

### Case Report

A previously healthy 35-year old gravida 3 para 1011 woman presented at 22 weeks gestation with symptoms of Covid-19 infection. Ten days prior to admission, she developed fever and cough, with acute worsening in the four days prior to admission including fever, malaise, nonproductive cough, diffuse myalgias, anorexia, nausea, and diarrhea. On the morning of presentation, the patient awoke with vaginal bleeding and abdominal pain. Her initial vital signs showed that she was afebrile with a pulse of 110, respiratory rate of 22 per minute and an oxygen saturation of 99% on room air. Her blood pressure was elevated to 150/100. Physical exam was notable for dark blood in the vaginal vault without cervical dilation. SARS-CoV-2 RNA was detected by RT-PCR in a nasopharyngeal swab obtained from the patient on admission.

Her past medical history was significant for psoriasis without current symptoms. The patient had a prior pregnancy that was complicated by term gestational hypertension which resolved with delivery. Her current antepartum course was notable for normal blood pressure and normal baseline preeclampsia evaluation.

The patient was admitted to labor and birth. Her chest x-ray was significant for a hazy opacity throughout the left lung. Transabdominal ultrasound revealed an active fetus, normal amniotic fluid volume, estimated fetal weight within expected range, and a posterior fundal placenta with a retroplacental clot concerning for placental abruption. Laboratory studies revealed elevated liver transaminases, profound thrombocytopenia, and increased urine protein consistent with preeclampsia, as well as prolonged partial thromboplastin time and decreased fibrinogen consistent with disseminated intravascular coagulation (Supplementary Table I). Blood smear revealed normochromic normocytic red cells with unremarkable morphology, atypical lymphocytes, suggestive of viral infection, and severe thrombocytopenia (Supplemental Figure S1). Rotational Thromboelastometry (ROTEM) demonstrated severe deficiencies in clot formation. The patient was resuscitated with 4 units of cryoprecipitate, 4 pools of packed platelets, 2 grams of tranexamic acid (TXA), 5 grams of fibrinogen concentrate, and 2 units of fresh frozen plasma. This resulted in the improvement of her coagulopathy, but the thrombocytopenia and elevated blood pressure persisted.

The combination of hypertension, proteinuria, elevated transaminases, and low platelets supported the diagnosis of severe preeclampsia, for which delivery is the definitive treatment. Multidisciplinary consultations were performed via telemedicine from maternal-fetal medicine, neonatology, and infectious disease. The patient opted for termination of the pre-viable pregnancy to reduce the risk of serious maternal morbidity or death, which was performed via dilation and evacuation under general endotracheal anesthesia. Intraoperative findings included a retroplacental clot and were otherwise unremarkable. On post-operative day 1, she was extubated and weaned to room air; however lymphopenia developed. Hydroxychloroquine was initiated as investigational treatment for Covid-19. Her coagulation markers improved and she was discharged to self-isolation on post-operative day three (Figure 1). Home blood pressure monitoring was initiated with close provider telemedicine support. An emergency room visit on post-operative day four was required to titrate antihypertensives. She consented to pathology examination and to release of tissue for research-related testing per our IRB-regulated protocol.

**Figure 1:**
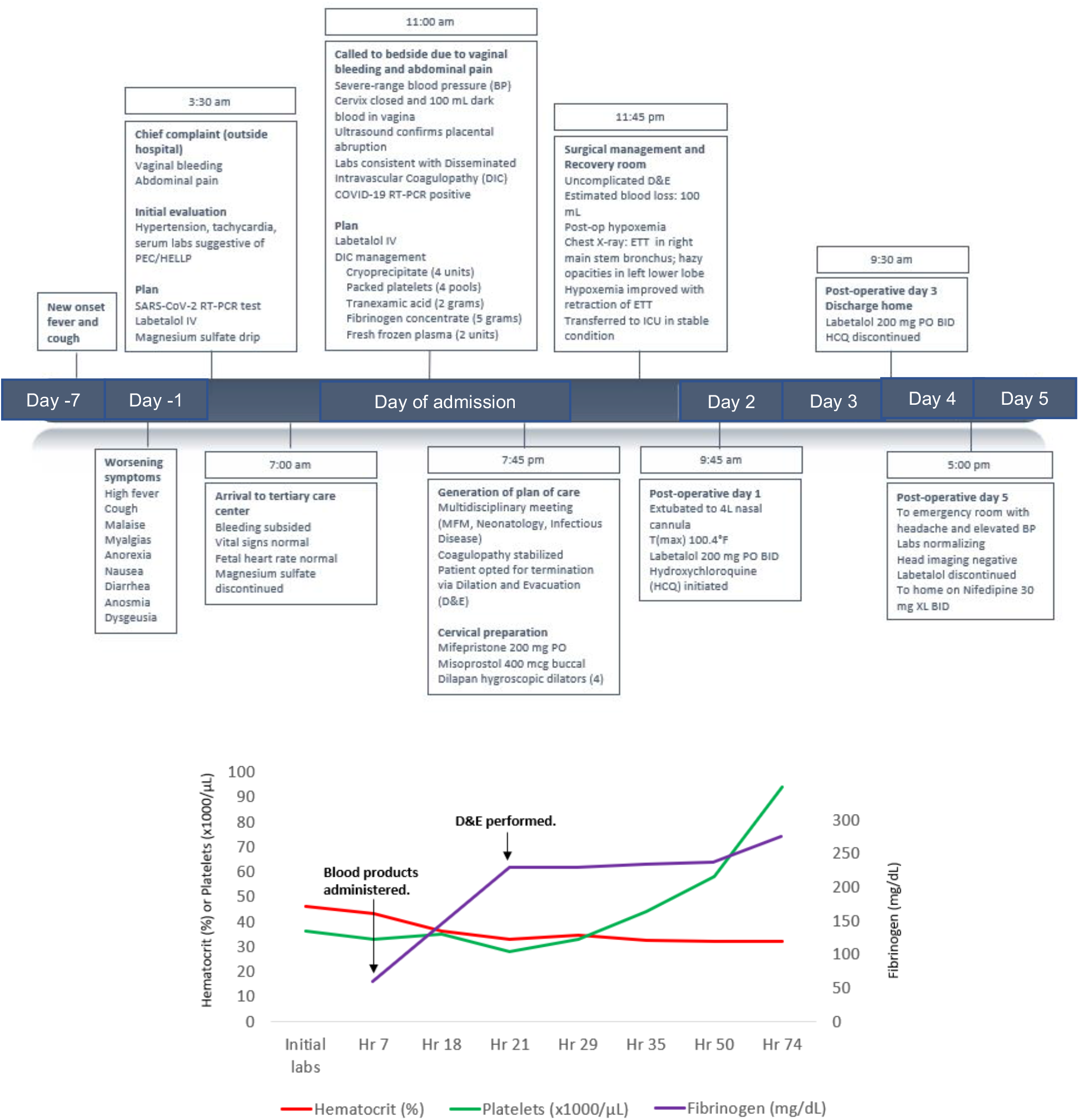
Case timeline. Clinical Course of 35 year old G3P1011 at 22 weeks gestation with Covid-19 associated with preeclampsia and placental abruption. Top panel: Timeline from onset of symptoms to immediate post-partum period including resuscitation products, hypertension management, and surgical preparation for dilation and evacuation, demonstrates Timeline from onset of symptoms to immediate post-partum period including resuscitation products, hypertension management, and surgical preparation for dilation and evacuation. Bottom panel: Patient’s platelet count (green line), and serum fibrinogen (purple line) throughout hospitalization. Major clinical events highlighted with arrows.

## RESULTS

### Microbiologic Investigation

Using the US CDC qRT-PCR assay, the placenta (3 x 10^7^ virus copies/mg) and umbilical cord (2 x 10^3^ virus copies/mg) were positive for SARS-CoV-2 RNA (Figure 2A). Fetal heart and lung tissues were also tested and met the human RNA control (RNase P) standards and were negative for virus RNA (Figure 2). Maternal samples were also collected post-operatively; and while the oral and nasal swabs were negative, the saliva and urine were still positive for SARS-CoV-2 (Figure 2A). Virus from the placenta was whole genome sequenced (Yale-050) and was phylogenetically similar to other SARS-CoV-2 detecting locally (Connecticut, USA) and abroad (Europe and Australia) (Figure 2B). In addition, the SARS-CoV-2 genome from the placenta did not contain any unique amino acid substitutions compared to other sequenced SARS-CoV-2.

**Figure 2:**
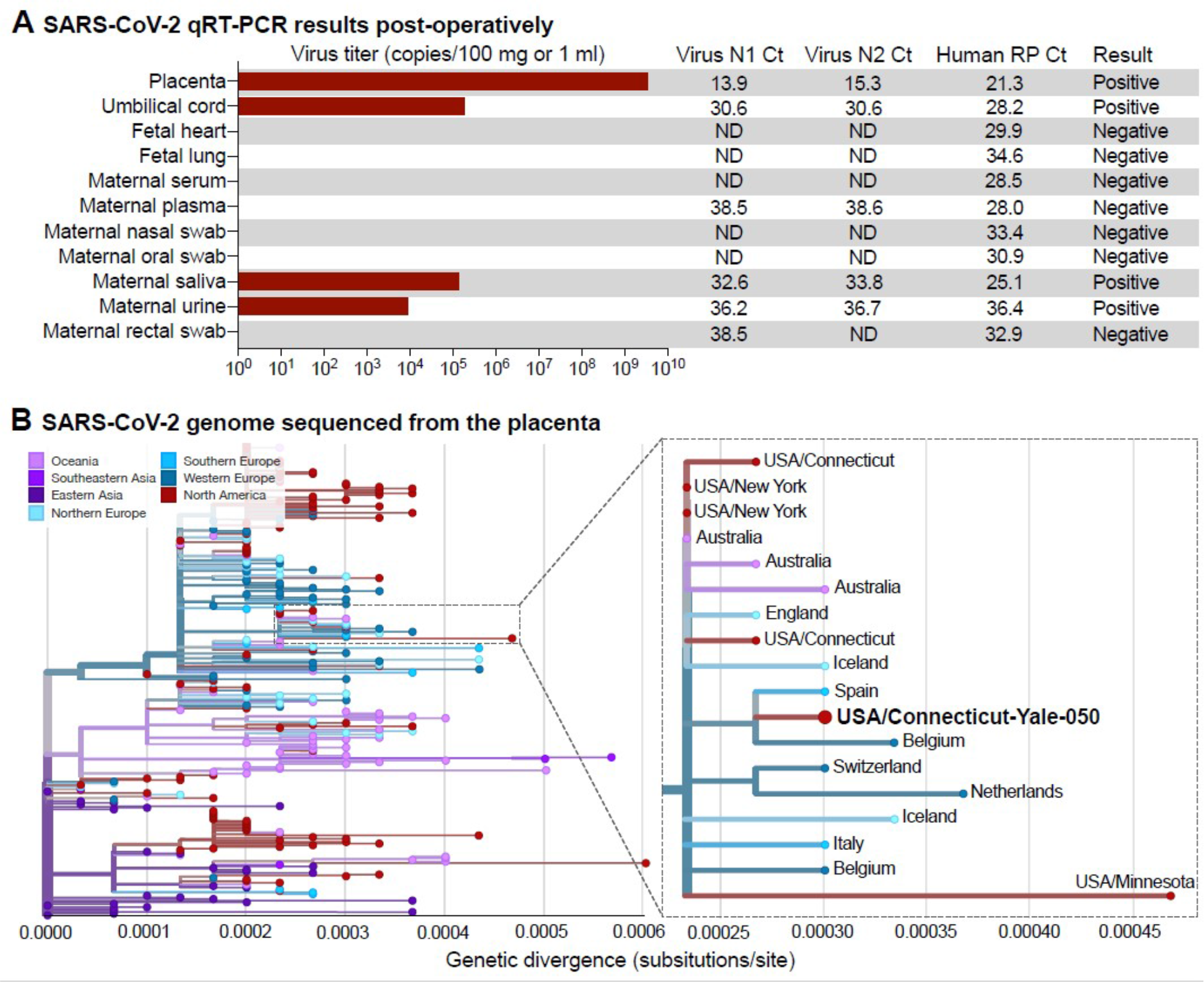
Examination of SARS-CoV-2 RNA in Maternal and Fetal Tissue. (**A**) SARS-CoV-2 qRT-PCR results of fetal and maternal samples using the CDC assay which consists of the N1 and N2 primers/probes targeting the coronavirus nucleocapsid and the RP primers/probe targeting human RNase P as an internal control. Cycle threshold (Ct) values from both N1 and N2 must be below 38 for the result to be positive, as internally validated. Virus titers shown as the average calculation from N1 and N2 Ct values. For the tissues, 80160 mg were used for extractions; and for the swabs in viral transport media and other liquid samples, 0.25-0.4 ml were used. (**B**) The SARS-CoV-2 genome sequenced from the infected placenta was combined with 289 other genomes available from GISAID from around the world. The phylogenetic tree was constructed using IQ-Tree within the Nextstrain Augur pipeline, and the results were visualized using Auspice. Genetic divergence is shown as substitutions per site from the root. An enlarged view of the 18 genomes in the clade that contains the SARS-CoV-2 genome sequenced from the placenta (USA/Connecticut-Yale-050). The clade is defined by 3 nucleotide substitutions, A28881A, G28882A, and G28883C, providing the equivalent of ~95% branch support. The consensus SARS-CoV-2 genome from the placenta (Yale-050) can be found using NCBI BioProject PRJNA614976 and the phylogenetic data can be visualized at: https://nextstrain.org/community/grubaughlab/CT-SARS-CoV-2/paper2. The acknowledgements for the sequences obtained from GISAID can be found at: https://github.com/grubaughlab/CT-SARS-CoV-2/tree/master/paper2.

### Serologic testing

Levels of anti-SARS-CoV-2 IgG and IgM antibodies in the case study patient were among the highest observed in 56 Covid-19^+^ patients admitted to Yale New Haven Hospital. Further quantification revealed end-point dilution titers of 1:1,600 for IgM and 1:25,600 for IgG (Supplemental Figure S2).

### Pathology

On gross examination the placenta showed a marginal adherent blood clot associated with a focal placental infarct supportive of the clinical diagnosis of abruption and was otherwise unremarkable. On histological examination the placenta was remarkable for the presence of diffuse perivillous fibrin and an inflammatory infiltrate composed of macrophages as well as T-lymphocytes, as demonstrated by immunohistochemistry for CD68 (Figure 3 A-C) and CD3 (Figure 3F). No dark pigment was noted in the intervillous space. Maternal vessels did not show features of decidual vasculopathy. The fetal organs were grossly and microscopically unremarkable (Supplemental Figure S3). SARS-CoV-2 localized predominantly to the syncytiotrophoblast cells of the placenta, as demonstrated by immunohistochemistry for the SARS-CoV-2 spike protein (Figure 3G-H), and by SARS-CoV-2 RNA *in situ* hybridization (Figure 3I).

**Figure 3:**
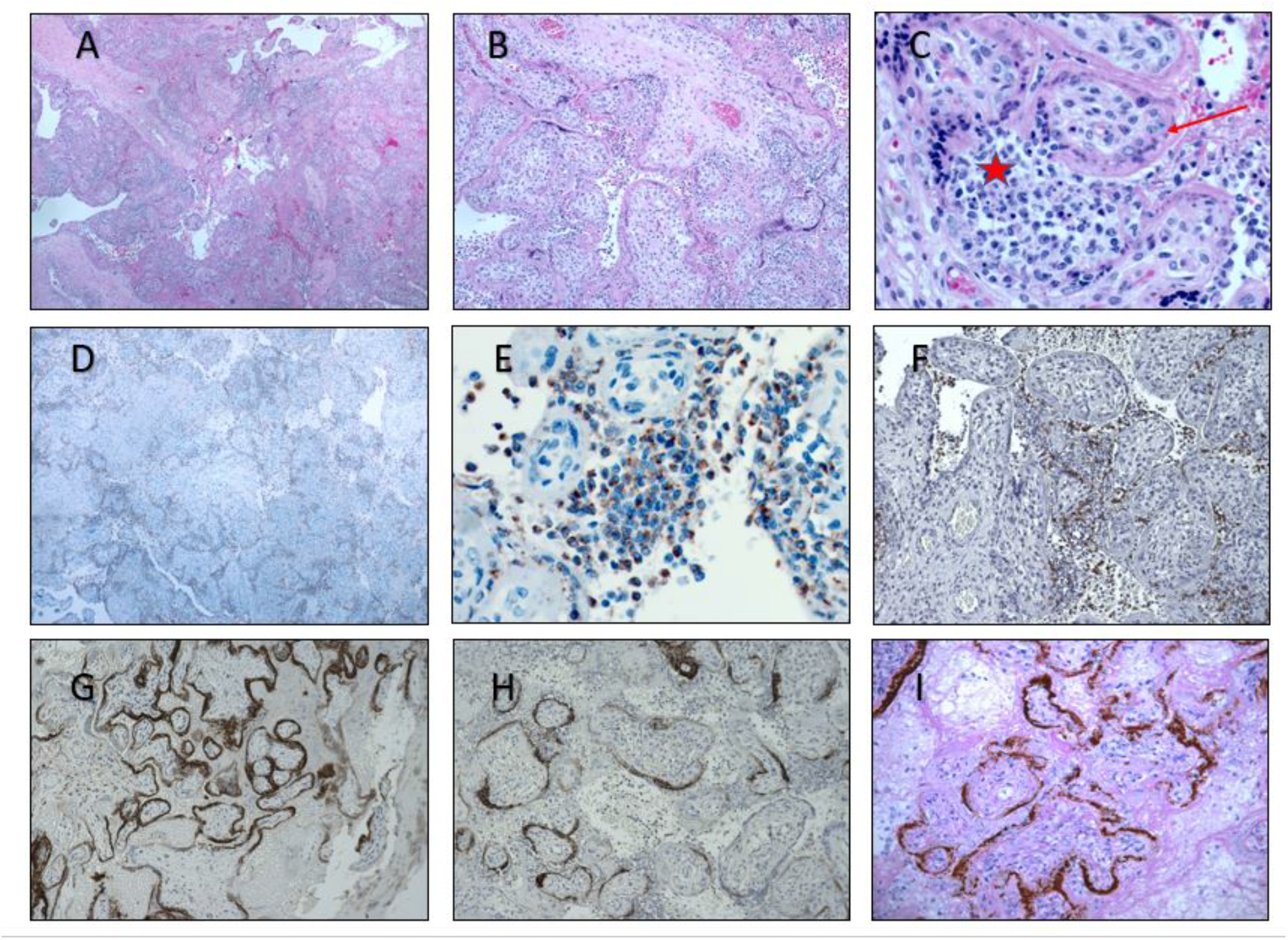
Histopathology of placenta. **A-C**. Section of placenta stained with Hematoxylin & Eosin showing histiocytic intervillositis (40X, 100X, 400X). **C**. Star indicates intervillous space infiltrated by immune cells. Arrows indicates perivillous fibrin. **D, E**. Immunohistochemical stain for CD68 showing the majority of intervillous inflammatory infiltrate positive (brown stain) for this macrophage marker (40X, 400X) **F**. Staining for CD3, a marker of T lymphocytes (100X). **G,H**. Immunohistochemical staining for SARS-CoV-2 spike protein, demonstrating viral localization predominantly in syncytiotrophoblast cells (50X, 100X). **I**. In situ analysis for the presence of SARS-CoV-2 RNA shows strong positive staining within the placenta.

### Electron Microscopy

Electron microscopic analysis of immersion-fixed placental tissue showed relatively well-preserved ultrastructure of the placenta (Figure 4). Analysis of placental region adjacent to the umbilical cord identified virus particles within the cytosol of placental cells. The size of these virus particles were 75-100nm in diameter, which is consistent with the size and appearance of SARS-CoV-2^12^.

**Figure 4:**
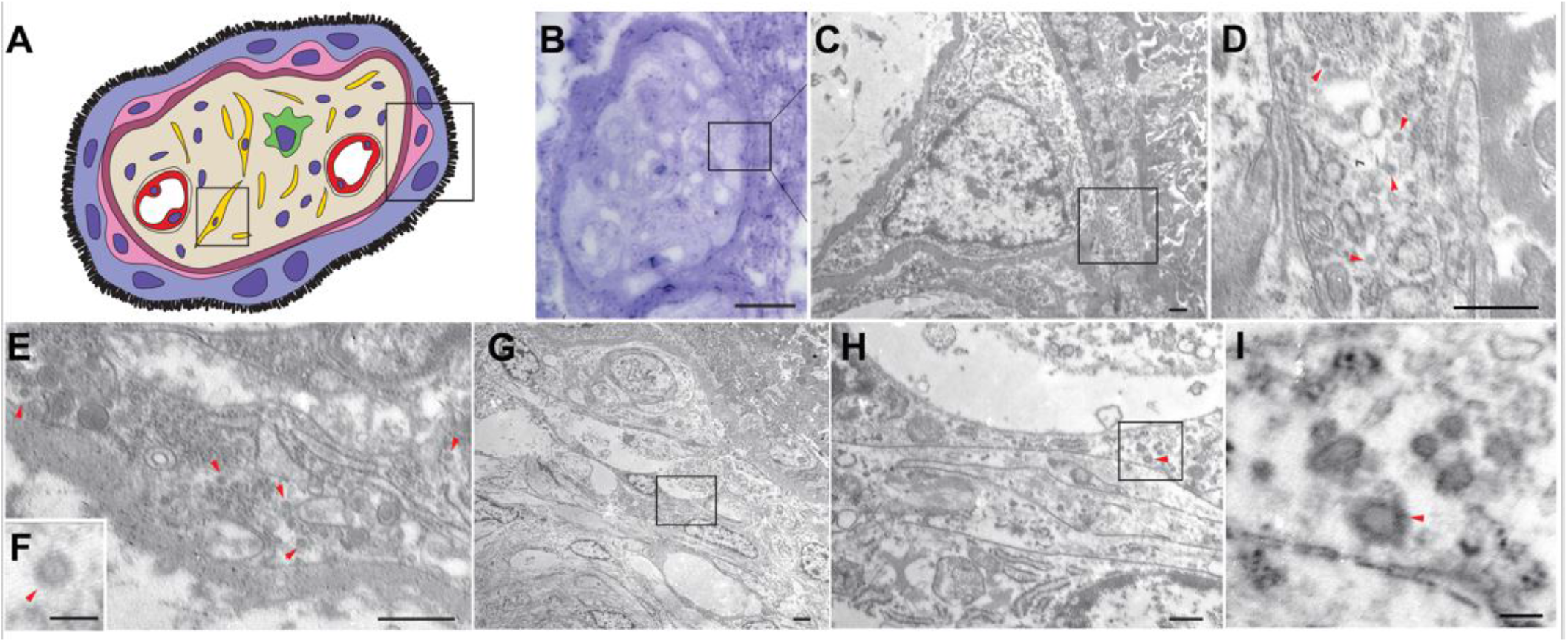
Electron microscopy images of coronavirus particles in different placental cell types. **A**. Schematic illustration of chorionic villi. Cytotrophoblast, syncytiotrophoblast and fibroblast are indicated with pink, purple and yellow, respectively. Right box indicates the location of cell on EM images of panels **C-F**. Left box indicates the origin of EM images of panels **G-I**. **B**. Toluidineblue staining of the section of which EM images on panels C-F are derived. Arrow originates from the that is depicted on panels **G-I**. Bar scale represents 25 nm. **C**. Cytotrophoblast (on the left with nucleus) and syncytiotrophoblast (on the right). Bar scale represents 500nm. **D**. Enlarged image of boxed area of panel **C**. Cytoplasm of a syncytiotrophoblast cells with virus particles (red arrowheads). Bare scale represents 500 nm **E**. Cytoplasm of a cytotrophoblast cell with virus particles indicated by red arrowheads. Bar scale represents 500 nm **F**. A representative high power magnification image of a virus particle. Bar scale represents 100 nm. **G**. Low power image of the chorionic villi stroma area. Bar scale represents 2 nm. **H**. Enlarged image of boxed area indicated on panel **G**. Cytoplasm of a fibroblast cell viral particles (red arrowheads). Bar scale represents 500 nm **I**. High-power image of boxed area of panel **H**. Red arrowhead indicates a virus particle. Bar scale represents 100 nm.

## DISCUSSION

This report describes a case of second-trimester Covid-19 associated with preeclampsia and SARS-CoV-2 infection of the placenta. The distinction between preeclampsia and Covid-19 is important, as it may have implications for the patient’s future pregnancy outcomes. While preeclampsia, placental abruption, and disseminated intravascular coagulopathy (DIC)—which commonly occur together in obstetric practice— account for many of the clinical and laboratory findings in this patient, the histopathological features and viral infection of the placenta suggest a prominent role for Covid-19 in this patient’s presentation. This is highlighted by the presence of high levels of SARS-CoV-2 and the invasion of intervillous macrophages (intervillositis) within the placenta. These findings suggest that Covid-19 may have contributed to placental inflammation that ultimately resulted in early-onset preeclampsia and worsening maternal disease.

Intervillositis, as seen in this case, describes a placental pathology consisting of fibrin deposits and mononuclear cell infiltration of the intervillous spaces. It is associated with high rates of miscarriage, fetal growth restriction, and severe early preeclampsia^13,14^. This entity is typically idiopathic or autoimmune in nature, but can also be seen in association with infections such as cytomegalovirus^15^ as well as malaria^16^. Massive fibrin deposition was also noted as a feature of placental pathology in pregnant women with SARS^17^ as well as malaria. In this case, it remains unknown whether Covid-19 precipitated intervillositis. However, the massive macrophage infiltration alongside fibrin deposition has recently been observed in lung tissue examined at autopsy from patients with severe Covid-19, raising the possibility of a common immunopathology leading to macrophage recruitment and activation causing tissue damage^18^. Further studies of placenta from women with Covid-19 may help address whether this is a histological feature associated with placental SARS-CoV-2 infection.

The pathophysiology of coagulopathy during Covid-19 remains unknown. A DIC-like coagulopathy has been reported in patients with Covid-19, and is associated with poor outcomes^19^. However, this patient’s thrombocytopenia and hypofibrinogenemia were more severe than what would have been expected from Covid-19 alone^19^. Both SARS-CoV-2 and hypertensive disorders of pregnancy are reported to reduce ACE2 activity, leading to increased tissue levels of angiotensin II^20-22^. The imbalance of the RAS system seen in Covid-19 patients may therefore contribute to hypertensive complications including preeclampsia in pregnant patients with Covid-19. We hypothesize that SARS-CoV2’s use of the ACE2 receptor could unmask HDP earlier in women who are predisposed to HDP and that Covid-19 causes significant placental pathology leading to an exacerbation in the severity of the presentation.

Importantly, there were no amino acid differences found in SARS-CoV-2 genome sequenced from the placenta compared to others sequenced from around the world (Figure 2), suggesting that placenta invasion is not a unique feature of this virus and it does not require adaptation. Rather, given the patient’s high serum titers of anti-SARS-CoV-2 spike protein antibodies, a potential mechanism to explain placental invasion in this case would be antibody dependent transcytosis mediated by the neonatal Fc receptor (FcRn), which has previously been observed for other viruses including CMV, HIV, and Zika^23-25^. Future studies are warranted to investigate protective versus pathological roles of humoral immunity in SARS-CoV-2 infection during pregnancy, and to assess for possible viral transmission to the fetus.

This report highlights a case of severe placental infection with SARS-CoV-2 which may have potentiated severe early-onset preeclampsia. Future efforts to correctly identify the diagnosis and underlying processes of Covid-19-associated hypertensive disorders in pregnancy are critical for directing patient care and counselling pregnant women during the pandemic.

## METHODS

### Microbiological investigation

RNA was extracted from homogenized placenta, umbilical cord, fetal lungs, heart kidney tissues (27-160 mg; stored in formalin) and maternal oral, nasal, and rectal swabs, saliva, urine, plasma, and serum postoperatively and tested for the presence of SARS-CoV-2 and human RNase P using the US CDC qRT-PCR assay as described^7^. The qRT-PCR results were confirmed via independent replicates. SARS-CoV-2 RNA from the placenta was sequenced using the portable MinION platform. The sequenced data was processed and used for phylogenetic analysis as described^8^.

### SARS-CoV-2 s1 spike protein IgM and IgG serology testing

ELISA assays for IgG and IgM antibodies towards SARS-CoV-2 were performed on maternal plasma as described by Amanat et al^9^. Initial screening of plasma samples from 56 Covid-19^+^ patients and 24 uninfected healthcare workers from the Yale New Haven Hospital was performed with a 1:50 dilution and endpoint titers were further tested for the case study patient using serial 4-fold dilutions beginning at 1:100. The cut-off was set with a confidence level of 99%, as described by Frey et al^10^. The endpoint titer is defined as the reciprocal of the highest dilution that gives a reading above the cut-off.

### Placenta and fetal organ examination

Placenta and fetal parts were immediately fixed in 10% buffered formalin. After three days of formalin fixation, placenta and fetal lung, heart, liver and kidney were sampled and embedded in paraffin. The placenta was sampled extensively with two sections of the cord and peripheral membranes and 10 full thickness sections of the placenta parenchyma which included both maternal and fetal side. Sections obtained were stained with hematoxylin and eosin (H&E). Immunohistochemical stains were performed on selected slides using antibodies for CD68 and CD3 using standard technique and for SARS-CoV-2 using the Monoclonal SARS-CoV-/SARS-CoV-2 spike antibody, clone 1A9 (Genetex; dilution 1:200). In situ hybridization for SARS-CoV-2 was performed with RNAScope (ACD, Neward, CA) using probes directed against SARS-CoV-2 on formalin fixed paraffin embedded tissue cut at thickness of 3 microns.^11^ Negative control probes (bacterial gene dapB) were also included to assess background signals as well as the positive control probes to the housekeeping gene peptidylprolyl isomerase B (PPIB). The ISH sections were counterstained using periodic acid-Schiff.

### Electron Microscopy

Tissues were fixed in 10% formaldehyde for 10 days. This solution was replaced by 4% paraformaldehyde and 0.3% glutaraldehyde for 20 hours. Tissue blocks then were processed for electron microscopy. Ultrathin sections were cut on a Leica Ultra-Microtome, collected on Formvar-coated single-slot grids, and analyzed with a Tecnai 12 Biotwin electron microscope (FEI).

### Human studies

This study was approved by the Yale Institutional Review Board. Written informed consent was obtained from the participant prior to inclusion in the study. The participant additionally agreed to inclusion in this report.

## Data Availability

data links in manuscript (sequencing)

## AUTHOR CONTRIBUTIONS

HH provided primary clinical care to the patient described in this case report and contributed to study conception and design, data collection and interpretation, literature searches, and writing the original manuscript. SF consented the patient for research, designed and oversaw all laboratory studies and interpretations, and aided in drafting the manuscript. HL designed and supervised all clinical care and study design, provided data interpretation, and contributed to both the drafting and clinical revisions of the manuscript. TH and YY contributed to electron microscopy. CV, JF, B, TA, AW, and NG contributed to viral detection, sequencing, and phylogenetics. AL, RM, RT, and CL contributed to histological studies. FL and AR contributed to antibody detection. CO, BG, PV, AC, JF, and CDC contributed to patient enrollment, specimen procurement, and tissue processing. AI, AK, TH, UD, KC, AG, GA, KF, HT, CP, and UR aided with study design, provided data interpretation, and contributed critical revisions of the manuscript. All authors reviewed and approved the final version of the manuscript. HH and SF share first authorship. HH appears first in the author list because she was primarily involved in the preparation and revisions of the manuscript until publication.

## ACKNOWLEDGEMENTS

We thank the patient in this case for her willingness to provide detailed medical and immunologic data, and Drs. Baxi, August and Webster for participating in this patient’s care.

---

**Supplemental Figure S1:**
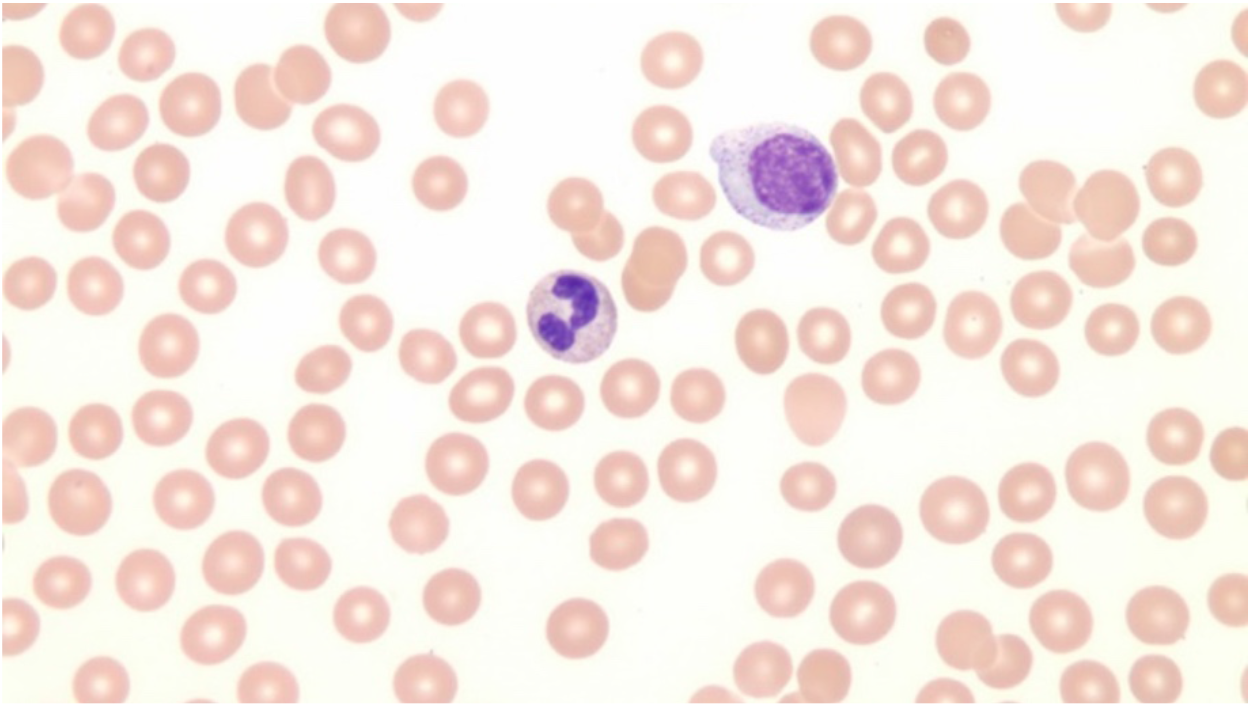
Patient blood smear. Blood smear shows normochromic normocytic red blood cells visualized without evidence of schistocytes or hemolysis. A polymorphic population of atypical lymphocytes were noted (not pictured) and severe thrombocytopenia without evidence of clumping.

**Supplemental Figure S2:**
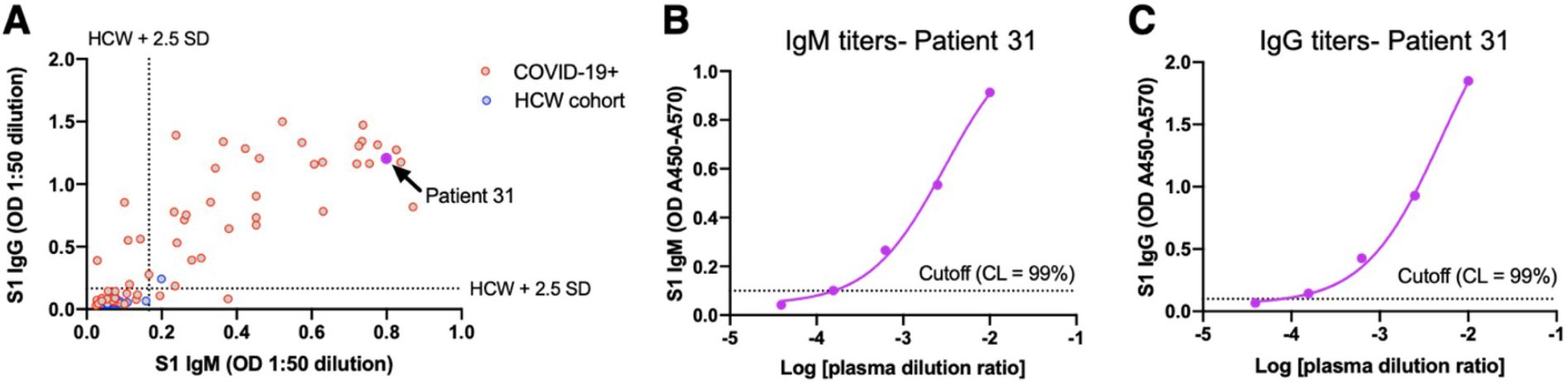
Quantification of anti-SARS-CoV-2 antibodies in maternal plasma. (A) Plot of ELISA optical densities (ODs) for IgM and IgG responses to the SARS-CoV-2 S1 spike protein from plasma samples of 56 COVID-19^+^ patients admitted to the Yale New Haven Hospital and 24 uninfected healthcare workers (HCW). All samples were assayed at a 1:50 dilution. The case study subject is indicated as patient 31. Dashed lines indicate the mean OD of the HCW cohort plus 2.5 standard deviations (SD). (B) Endpoint dilution titers of IgM and (C) IgG for the case study patient. The 99% confidence cut-off limit is depicted with a dashed line. The endpoint titer for IgM was 1:1,600 and 1:25,600 for IgG.

**Supplemental Figure S3:**
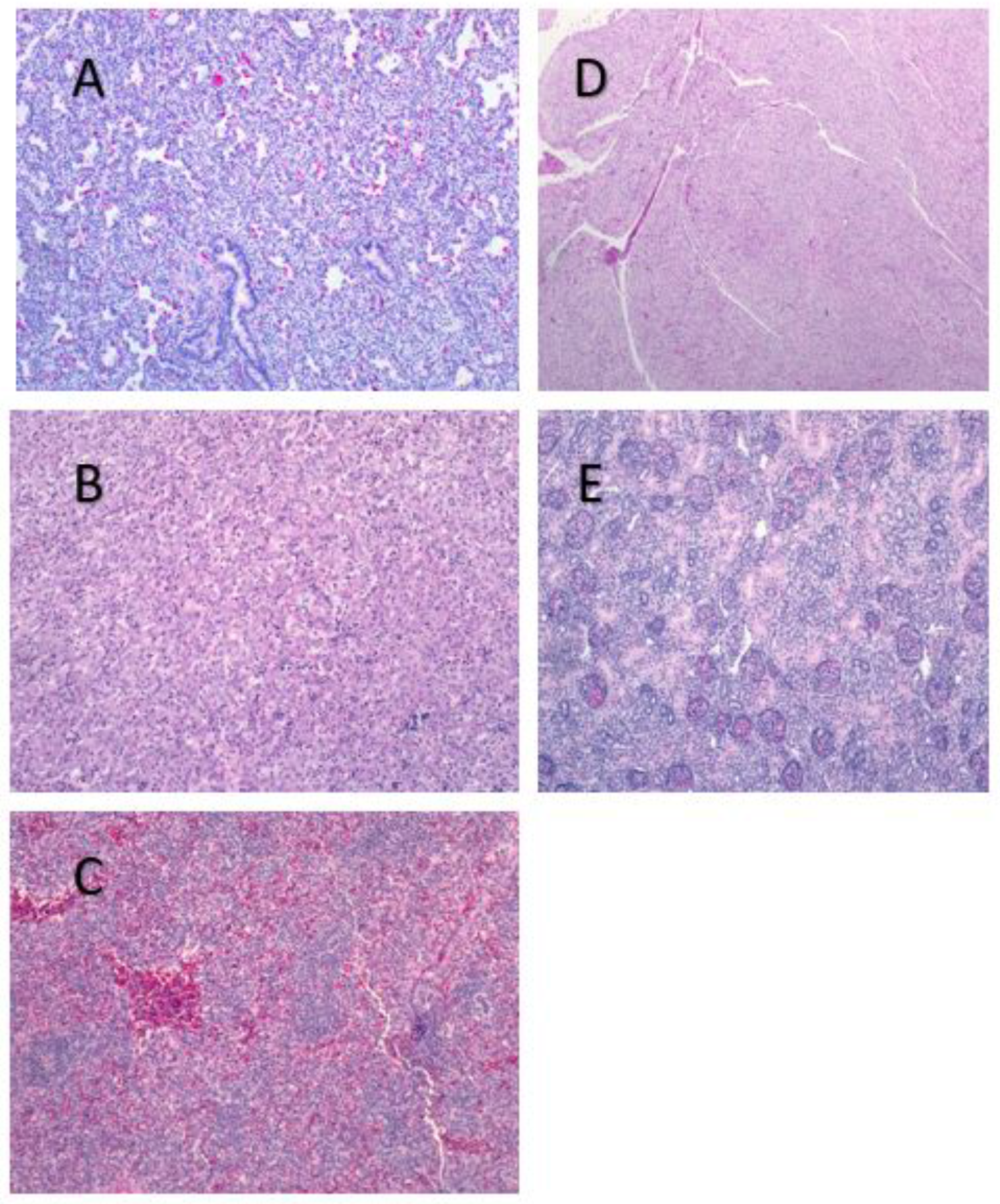
Fetal organ histology. H&E stain of fetal (A) lung, (B) myocardium, (C) liver, (D) kidney, and (E) spleen show normal histology without inflammatory changes.

**Supplementary Table I:**
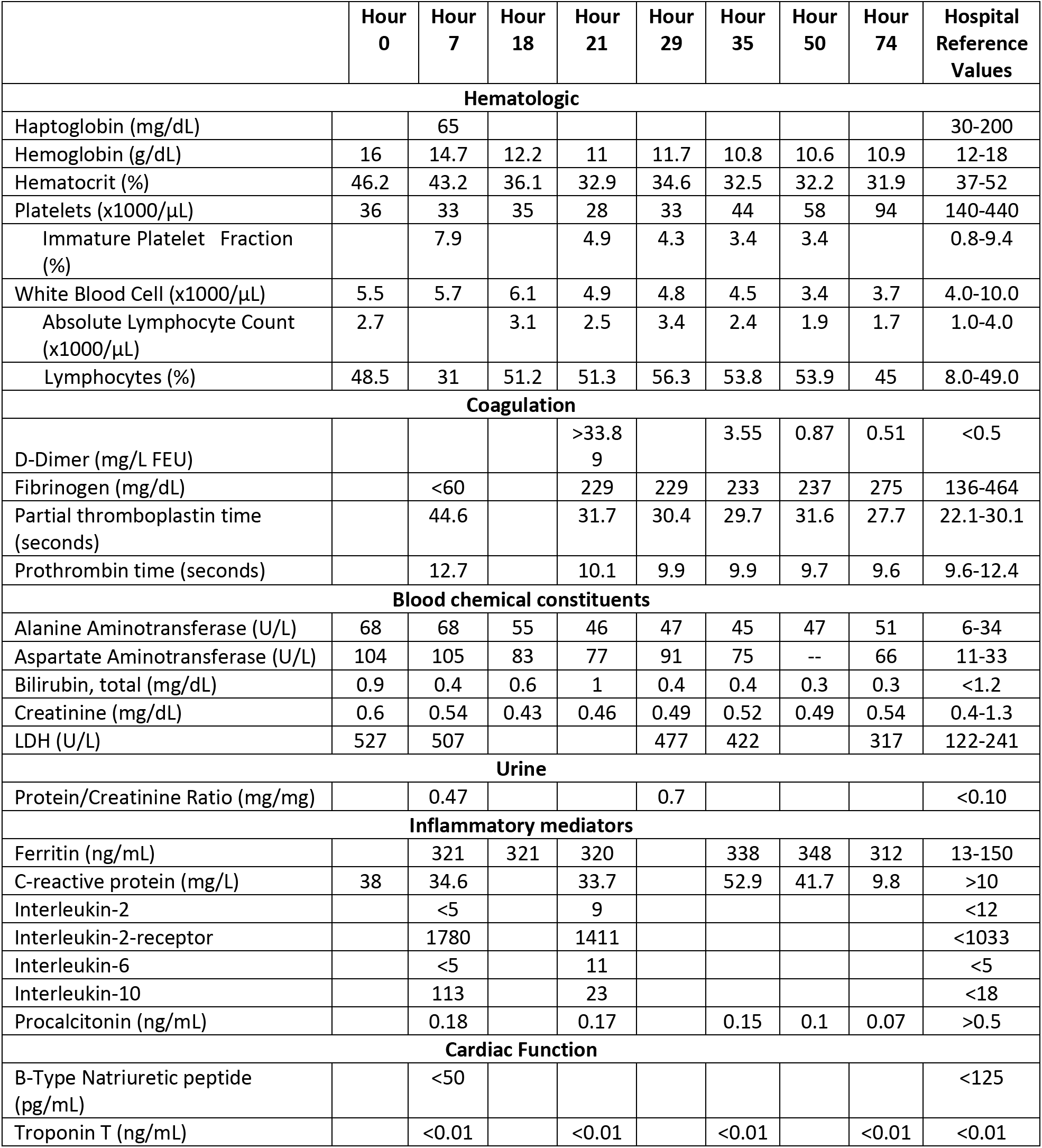
Patient’s serial laboratory results from presentation to post-operative day 3.

| Table 1. Serial Laboratory Results |  |  |  |  |  |  |  |  |  |
| --- | --- | --- | --- | --- | --- | --- | --- | --- | --- |
|  | Hour<br>0 | Hour<br>7 | Hour<br>18 | Hour<br>21 | Hour<br>29 | Hour<br>35 | Hour<br>50 | Hour<br>74 | Hospital<br>Reference<br>Values |
| **Hematologic** |  |  |  |  |  |  |  |  |  |
| Haptoglobin (mg/dL) |  | 65 |  |  |  |  |  |  | 30-200 |
| Hemoglobin (g/dL) | 16 | 14.7 | 12.2 | 11 | 11.7 | 10.8 | 10.6 | 10.9 | 12-18 |
| Hematocrit (%) | 46.2 | 43.2 | 36.1 | 32.9 | 34.6 | 32.5 | 32.2 | 31.9 | 37-52 |
| Platelets (x1000/μL) | 36 | 33 | 35 | 28 | 33 | 44 | 58 | 94 | 140-440 |
| Immature Platelet Fraction (%) |  | 7.9 |  | 4.9 | 4.3 | 3.4 | 3.4 |  | 0.8-9.4 |
| White Blood Cell (x1000/μL) | 5.5 | 5.7 | 6.1 | 4.9 | 4.8 | 4.5 | 3.4 | 3.7 | 4.0-10.0 |
| Absolute Lymphocyte Count (x1000/μL) | 2.7 | | 3.1 | 2.5 | 3.4 | 2.4 | 1.9 | 1.7 | 1.0-4.0 |
| Lymphocytes (%) | 48.5 | 31 | 51.2 | 51.3 | 56.3 | 53.8 | 53.9 | 45 | 8.0-49.0 |
| **Coagulation** |  |  |  |  |  |  |  |  |  |
| D-Dimer (mg/L FEU) |  |  |  | >33.8<br>9 |  | 3.55 | 0.87 | 0.51 | <0.5 |
| Fibrinogen (mg/dL) |  | <60 |  | 229 | 229 | 233 | 237 | 275 | 136-464 |
| Partial thromboplastin time (seconds) |  | 44.6 |  | 31.7 | 30.4 | 29.7 | 31.6 | 27.7 | 22.1-30.1 |
| Prothrombin time (seconds) |  | 12.7 |  | 10.1 | 9.9 | 9.9 | 9.7 | 9.6 | 9.6-12.4 |
| **Blood chemical constituents** |  |  |  |  |  |  |  |  |  |
| Alanine Aminotransferase (U/L) | 68 | 68 | 55 | 46 | 47 | 45 | 47 | 51 | 6-34 |
| Aspartate Aminotransferase (U/L) | 104 | 105 | 83 | 77 | 91 | 75 | -- | 66 | 11-33 |
| Bilirubin, total (mg/dL) | 0.9 | 0.4 | 0.6 | 1 | 0.4 | 0.4 | 0.3 | 0.3 | <1.2 |
| Creatinine (mg/dL) | 0.6 | 0.54 | 0.43 | 0.46 | 0.49 | 0.52 | 0.49 | 0.54 | 0.4-1.3 |
| LDH (U/L) | 527 | 507 |  |  | 477 | 422 |  | 317 | 122-241 |
| **Urine** |  |  |  |  |  |  |  |  |  |
| Protein/Creatinine Ratio (mg/mg) |  | 0.47 |  |  | 0.7 |  |  |  | <0.10 |
| **Inflammatory mediators** |  |  |  |  |  |  |  |  |  |
| Ferritin (ng/mL) |  | 321 | 321 | 320 |  | 338 | 348 | 312 | 13-150 |
| C-reactive protein (mg/L) | 38 | 34.6 |  | 33.7 |  | 52.9 | 41.7 | 9.8 | >10 |
| Interleukin-2 |  | <5 |  | 9 |  |  |  |  | <12 |
| Interleukin-2-receptor |  | 1780 |  | 1411 |  |  |  |  | <1033 |
| Interleukin-6 |  | <5 |  | 11 |  |  |  |  | <5 |
| Interleukin-10 |  | 113 |  | 23 |  |  |  |  | <18 |
| Procalcitonin (ng/mL) |  | 0.18 |  | 0.17 |  | 0.15 | 0.1 | 0.07 | >0.5 |
| **Cardiac Function** |  |  |  |  |  |  |  |  |  |
| B-Type Natriuretic peptide (pg/mL) |  | <50 |  |  |  |  |  |  | <125 |
| Troponin T (ng/mL) |  | <0.01 |  | <0.01 |  | <0.01 |  | <0.01 | <0.01 |

